# A Methodology Framework for Analyzing Health Misinformation to Develop Inoculation Intervention Using Large Language Models: A case study on covid-19

**DOI:** 10.1101/2025.05.22.25327931

**Authors:** Samira Malek, Christopher Griffin, Robert Fraleigh, Robert P. Lennon, Vishal Monga, Lijiang Shen

## Abstract

**Background:** The rapid growth of social media as an information channel has enabled the swift spread of inaccurate or false health information, significantly impacting public health. This widespread dissemination of misinformation has caused confusion, eroded trust in health authorities, led to noncompliance with health guidelines, and encouraged risky health behaviors. Understanding the dynamics of misinformation on social media is essential for devising effective public health communication strategies.

**Objective:** This study aims to present a comprehensive and automated approach that leverages Large Language Models (LLMs) and Machine Learning (ML) techniques to detect misinformation on social media, uncover the underlying causes and themes, and generate refutation arguments, facilitating control of its spread and promoting public health outcomes by inoculating people against health misinformation.

**Methods:** We use two datasets to train three LLMs, namely BERT, T5, and GPT-2, to classify documents into two categories: misinformation and non-misinformation. Additionally, we employ a separate dataset to identify misinformation topics. To analyze these topics, we apply three topic modeling algorithms—Latent Dirichlet Allocation (LDA), Top2Vec, and BERTopic—and selected the optimal model based on performance evaluated across three metrics. Using a prompting approach, we extract sentence-level representations for the topics to uncover their underlying themes. Finally, we design a prompt text capable of identifying misinformation themes effectively.

**Results:** The trained BERT model demonstrated exceptional performance, achieving 98% accuracy in classifying misinformation and non-misinformation, with a 44% reduction in false positive rates for AI-generated misinformation. Among the three topic modeling approaches employed, BERTopic outperformed the others, achieving the highest metrics with a Coherence Value (CV) of 0.41, Normalized Pointwise Mutual Information (NPMI) of -0.086, and Inverted RBO (IRBO) of 0.99. To address the issue of unclassified documents, we developed an algorithm to assign each document to its closest topic. Additionally, we proposed a novel method using prompt engineering to generate sentence-level representations for each topic, achieving a 99.6% approval rate as “appropriate” or “somewhat appropriate” by three independent raters. We further designed a prompt text to identify themes of misinformation topics and developed another prompt capable of detecting misinformation themes with 80% accuracy.

**Conclusions:** This study presents a comprehensive and automated approach to addressing health misinformation on social media using advanced machine learning and natural language processing techniques. By leveraging large language models (LLMs) and prompt engineering, the system effectively detects misinformation, identifies underlying themes, and provides explanatory responses to combat its spread.

(Journal of Medical Internet Research) doi:

## Introduction

Misinformation and inaccurate beliefs and knowledge about health can substantially undermine well-being by fueling confusion, eroding trust in reliable medical advice, and prompting risky behaviors such as rejecting vaccines, turning to scientifically unproven home remedies, or neglecting protective measures amid clear dangers (1–8). These inaccuracies often circulate rapidly via social media, exploiting emotional narratives that overshadow fact-based content and leading individuals to question the legitimacy of evidence-based interventions (5,8–11). Repeated exposure to misinformation reduces health literacy and can reinforce people’s belief in falsehoods, making them more likely to view credible health authorities with skepticism (12–14). As a result, misinformation weakens the success of prevention and treatment strategies, paving the way for heightened disease transmission, avoidable complications, and deteriorating outcomes at both individual and community levels (15–18).

An illustration comes from the COVID-19 pandemic, which saw an unprecedented surge of misinformation and conspiracy theories—labeled an “infodemic” by the World Health Organization (1,19). False remedies, unverified claims on the origins of the virus, and politicized narratives about preventive measures severely hampered containment efforts (20–22). While proven strategies such as mask-wearing, vaccination, and physical distancing were promoted by scientific authorities, social media rumors cast doubt on vaccine safety and the reality of the virus itself, discouraging people from getting vaccinated or seeking appropriate medical care (2,23–25). This breakdown in adherence prolonged outbreaks, overloaded health infrastructures, and ultimately jeopardized global health and economic stability (26).

A parallel can be drawn from discussions around the human papillomavirus (HPV) vaccine, which has proven crucial in preventing various HPV-related cancers, including cervical cancer that claims thousands of lives each year (27–31). Widespread misinformation about adverse effects and conspiracies regarding its necessity led to a significant portion of unvaccinated adolescents, heightening the likelihood of HPV infection and future malignancies (32). This trend not only increased the burden on public health systems but also underscored the power of misinformation to undermine trust in legitimate medical counsel.

In recent years, social media has become a central and highly accessible source of information for millions of users worldwide(33). However, its ability to rapidly disseminate content—including unfounded claims—creates fertile ground for large-scale propagation of misinformation. Given the sheer volume of posts manual monitoring and analysis of such content are impractical (34,35). Consequently, developing and employing automated, data-driven methods to understand and manage the dynamics of digital misinformation is essential for preserving accurate information and safeguarding public trust.

In this study, we introduce the Misinformation Detection and Inoculation Process (MDIP) for analyzing health misinformation, along with a complementary Misinformation Detection and Inoculation System (MDIS) that generates refutation arguments (inoculation) to help prevent its spread on social media and enhance public health awareness (36). To achieve this, we leverage a Large Language Model (LLM) to detect misinformation effectively. Furthermore, we demonstrate that enriching datasets improves the detection of misinformation generated by both humans and AI. Recent advances in LLMs, such as ChatGPT, have enabled the generation of increasingly sophisticated misinformation, which poses challenges for traditional machine learning (ML) methods in distinguishing AI-generated misinformation (37,38). While prior research has highlighted the effectiveness of Deep Learning (DL) methods in classifying health-related misinformation, these efforts have predominantly focused on content generated by humans (39,40). Moreover, our proposed process generates sentence-level descriptions of misinformation topics, eliminating the need for manual interpretation. However, prior approaches relied on ML-based methods that produced word-level topic representations, which required manual interpretation to form coherent sentence-level topics—introducing potential human errors and subjective biases (1,23). Similar challenges arise in other applications, such as optimizing models in industries where manual calibration of parameters can lead to inefficiencies and errors. Recent research has demonstrated that data-driven models can enhance predictive accuracy and automate decision-making (41–43), reducing human intervention in systems that rely on complex data streams (44–49). Inspired by these advances, our process generates sentence-level descriptions of misinformation topics, eliminating the need for manual interpretation. Additionally, we introduce an algorithm to assign documents to the most relevant topics. This addresses the limitation of many ML-based topic modeling algorithms, which often leave some documents unclassified. Our process also identifies overarching themes of misinformation topics automatically, providing a high-level understanding of the underlying reasons for misinformation categorization.

## Methods

In this study, we propose the Misinformation Detection and Inoculation Process (MDIP), a comprehensive framework designed to analyze the dynamics of misinformation automatically and develop a Misinformation Detection and Inoculation System (MDIS). The MDIP framework is structured into four interconnected sections, each addressing a critical aspect of misinformation management:

- **Detect Misinformation:** This section uses large language models (LLMs) to classify text documents as misinformation or non-misinformation, providing the foundation for identifying and analyzing false narratives.
- **Misinformation Topics:** Here, topic modeling algorithms are applied to uncover the key topics within misinformation datasets. This step helps to categorize misinformation into specific subject areas, enabling a better understanding of its thematic structure.
- **Topic Descriptions:** This section enhances interpretability by generating sentence-level representations for each topic, moving beyond traditional word-level outputs. These descriptive summaries provide meaningful context for understanding the essence of each topic.
- **Provide Refutation:** In the final step, the system generates clear and contextually relevant refutation arguments for the identified misinformation themes. These arguments are designed to counter false narratives, improve public understanding, and mitigate the spread of misinformation.

By integrating these components, MDIP enables the development of MDIS, an intelligent and automated system capable of detecting misinformation, identifying its themes, and delivering refutations to combat its impact on public health.

### Detect Misinformation

Misinformation detection in text documents has become a critical area of research due to the growing prevalence of misleading or false information online. To address this challenge, we employ classifiers based on LLMs. These LLMs are trained to categorize text into two classes: misinformation and non-misinformation as shown in the first part of Figure 1.

**Figure 1.**
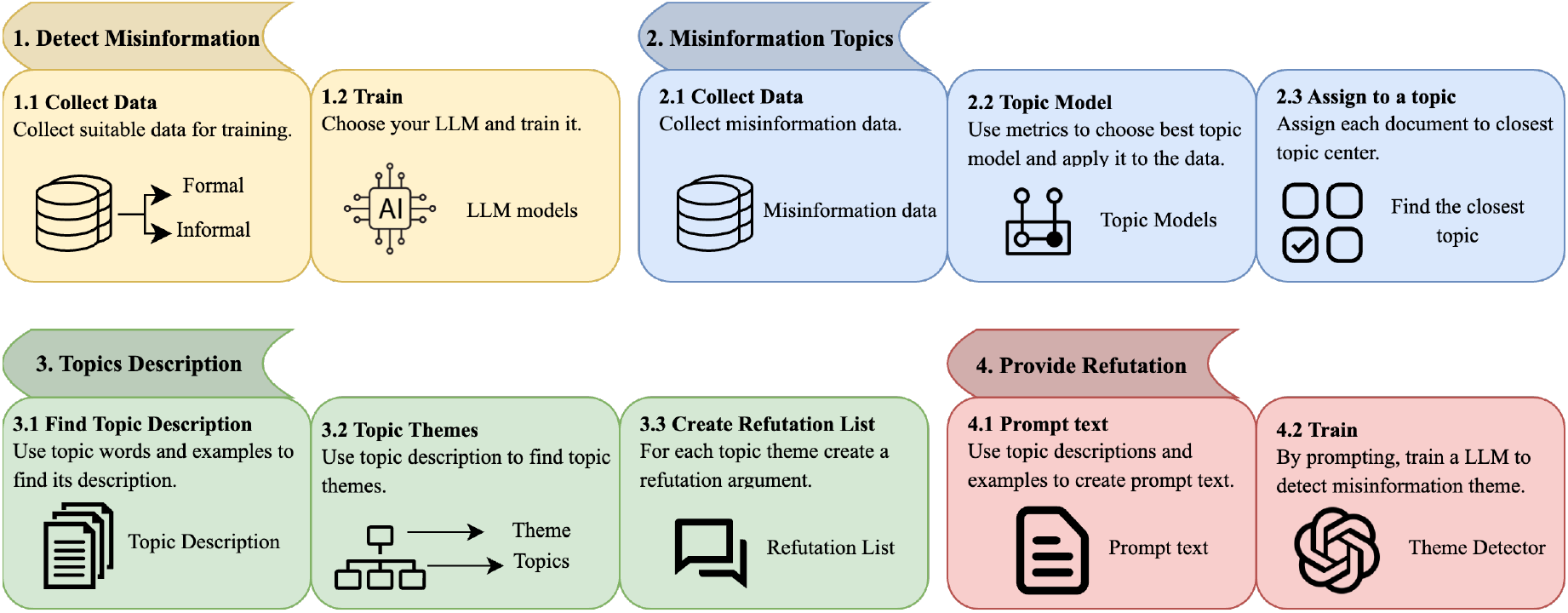
Overview of the Misinformation Detection and Inoculation Process (MDIP), consisting of four components: 1. Detect Misinformation, 2. Misinformation Topics, 3. Topic Descriptions, and 4. Provide Refutation.

Our classifier was trained using two complementary datasets, each providing diverse linguistic characteristics to enhance performance in detecting misinformation. The first dataset, the AAAI 2021 Competition Dataset (50), consists of misinformation sourced from social media platforms such as Facebook and X (formerly known as Twitter). This dataset reflects the informal, conversational style of social media, characterized by casual tone, non-standard grammar, and the use of slang. The second dataset, COVID_19FNIR (51), includes misinformation presented in formal, structured language, offering a stark contrast to the informal nature of the first dataset. By incorporating these two datasets, we trained three different LLMs to detect misinformation effectively across a wide spectrum of communication styles. The blend of informal and formal language enabled the model to better generalize, achieving improved accuracy and robustness in identifying misinformation, whether generated by humans or artificial intelligence (AI).

Traditionally, researchers collect human-written data from social media platforms like Twitter and Facebook, label it as misinformation or non-misinformation, and then train a Deep Neural Network (DNN) to classify such documents (39,40,52,53). However, recent studies have demonstrated that DNNs trained exclusively on human-written datasets exhibit weaker accuracy in detecting AI-generated misinformation compared to human-written misinformation. This discrepancy arises because AI-generated misinformation often adopts formal language styles similar to accurate information shared by credible sources like the Word Health Organization (WHO) and Centers for Disease Control and Prevention (CDC) on official social media accounts (37,38).

In this research, by combining a dataset with formal language and another with informal language, we demonstrate that LLMs achieve superior accuracy in detecting AI-generated misinformation. This approach ensures better generalization and robustness, bridging the gap in identifying misinformation across diverse linguistic styles.

### Misinformation Topics

As outlined in the second section of Figure 1, our approach involves three key steps. First, we collect misinformation data. Next, we select and compare topic modeling algorithms based on specific features and metrics to identify the most effective model. Finally, we design an algorithm that assigns topics to new or unclassified documents.

To identify misinformation topics, we used one of the largest datasets of verified COVID-19 claims, the IFCN dataset, which has been extensively used in related research (54–56). We applied three topic modeling algorithms—Latent Dirichlet Allocation (LDA) (57), Top2Vec (58,59), and BERTopic (59,60)—to analyze this dataset.

To evaluate and compare the performance of these algorithms, we selected three metrics: Coherence Value (CV) (61), Normalized Pointwise Mutual Information (NPMI) (62), and Inverse Rank-Biased Overlap (IRBO) (63). CV and NPMI measure the coherence of the topics, ensuring they are logically consistent and meaningful. IRBO, on the other hand, evaluates the diversity of the topics generated by the model, which is crucial for ensuring broad coverage of the dataset’s content. Since our focus is on misinformation within health-related social media data, coherence and diversity are particularly important to ensure topics are both interpretable and representative.

After selecting the best-performing topic model, we developed an algorithm to address the issue of unclassified documents. This algorithm assigns topics to new or previously unassigned documents, ensuring comprehensive topic coverage and improved usability of the model for real-world applications.

### Topic Description

Topic modeling algorithms typically produce word-level representations for each topic. While these representations provide insight into the most relevant words associated with a topic, they often lack the semantic depth necessary to precisely identify the specific topic within a document. This limitation arises because word-level outputs fail to capture the context and relationships between words that define the overarching theme of a topic (52).

Recent advancements in large language models (LLMs) have demonstrated their ability to generate high-quality, contextually relevant outputs with minimal or zero additional training by designing carefully crafted inputs—referred to as prompt engineering (64). Leveraging this capability, we address the limitations of word-level representations by employing prompt engineering techniques to generate sentence-level representations for each topic. These sentence-level representations capture the context and essence of the topic, enabling a more accurate and interpretable understanding of the document content.

Subsequently, these sentence-level representations are used to identify and articulate the overarching themes of the topics, also at the sentence level. This approach provides a more comprehensive view of the thematic structure within the document corpus.

Finally, recognizing that all documents within the dataset share a common underlying reason for being classified as misinformation, we develop a tailored response list for each topic theme. The third section of Figure 1 illustrates these three steps.

### Provide Refutation

In the final step of our proposed method, as illustrated in the final part of Figure 1, we identify the overarching theme of misinformation and provide a corresponding response from a pre-constructed response list. This response list is developed in the preceding step based on the identified themes.

To determine the themes of misinformation, we use prompt engineering techniques. By designing carefully crafted and contextually appropriate prompt text, we effectively extract the underlying themes associated with misinformation. This approach allows us to translate complex word-level or sentence-level representations into meaningful thematic insights.

By identifying misinformation themes and providing precise, theme-based responses, our method aims to enhance public health knowledge and reduce the spread of misinformation. This proactive approach not only mitigates the risks associated with false or misleading information but also fosters a more informed and resilient society.

### Proposed System

Following the completion of four foundational steps—1) Detecting misinformation, 2) Identifying misinformation topics, 3) Describing topics, and 4) Providing Refutations—we develop our comprehensive Misinformation Detection and Inoculation System (MDIS), which consists of three key components.

1. **Detection of Misinformation**: The system begins by determining whether a given document is misinformation.
2. **Identification of Misinformation Themes**: If the document is classified as misinformation, the system analyzes its content to identify the underlying misinformation themes. This process involves extracting thematic representations that provide a clearer understanding of the document’s misleading aspects.
3. **Providing Refutations:** Finally, the system generates a detailed refutation argument for the identified misinformation themes. These arguments are derived from a pre-designed response list tailored to address specific misinformation themes effectively.

All three components of the system are demonstrated with a practical example, as illustrated in Figure 2. This example highlights how the system operates cohesively to detect misinformation, uncover its thematic structure, and deliver accurate refutations, ultimately contributing to a more informed and resilient public. While this approach provides a structured framework, it has not been implemented as a real-time system or integrated into public health platforms.

**Figure 2.**
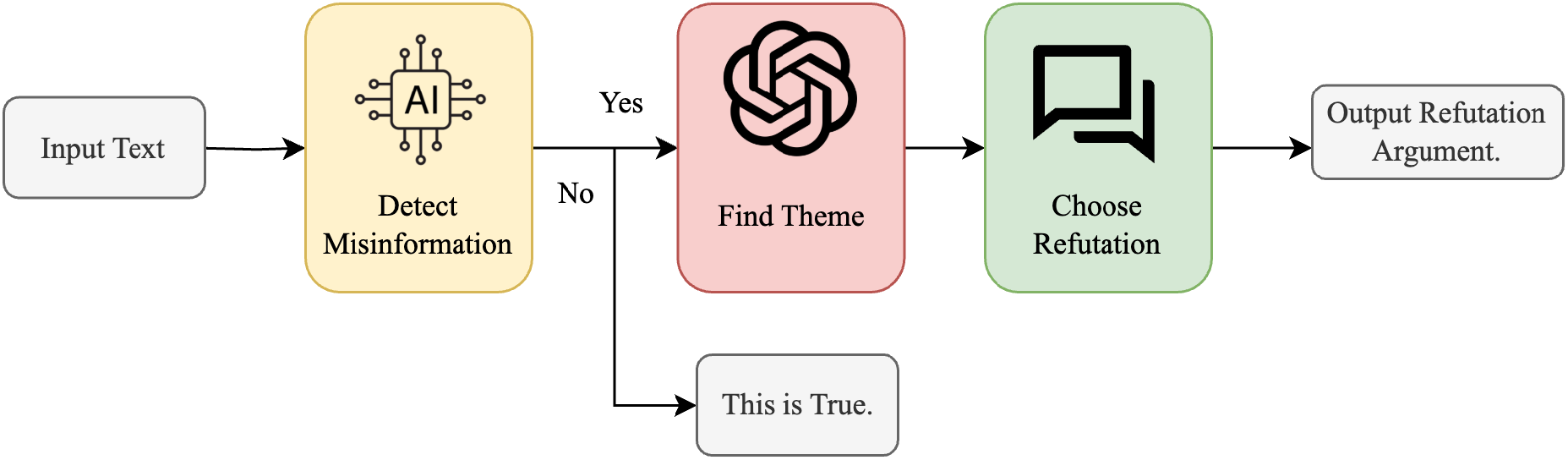
Misinformation Detection and Inoculation System (MDIS).

## Results

### Text Classification

Due to the exceptional performance of Large Language Models (LLMs) across a wide range of artificial intelligence tasks, we leveraged three prominent LLMs to fine-tune them for COVID-19 text classification. These models—BERT, GPT-2, and T5-base—are renowned for their ability to understand and process natural language with high accuracy and contextual awareness.

BERT (Bidirectional Encoder Representations from Transformers) is particularly effective in handling text classification tasks due to its bidirectional context understanding, which allows it to capture nuanced language patterns (65). GPT-2 (Generative Pre-trained Transformer 2) excels in text generation and classification by leveraging its autoregressive architecture to predict sequences in a given context (66). Lastly, T5-base (Text-to-Text Transfer Transformer) operates under a unified framework that reformulates all NLP tasks as a text-to-text problem, making it versatile and effective across various domains (67).

To conduct this study, we combined the AAAI 2021 competition dataset with the COVID-19FNIR dataset. The data was split into training, testing, and validation sets with proportions of 67%, 17%, and 16%, respectively.

Table 1 shows the evaluation metrics, including Accuracy, F1-Score, Recall, and Precision, for all three models on the test dataset. Among these, BERT achieved the highest performance, with an impressive accuracy of 98% on the test data. This superior result highlights BERT’s ability to handle complex linguistic structures and its effectiveness in fine-tuning for domain-specific tasks like COVID-19 text classification.

**Table 1.**
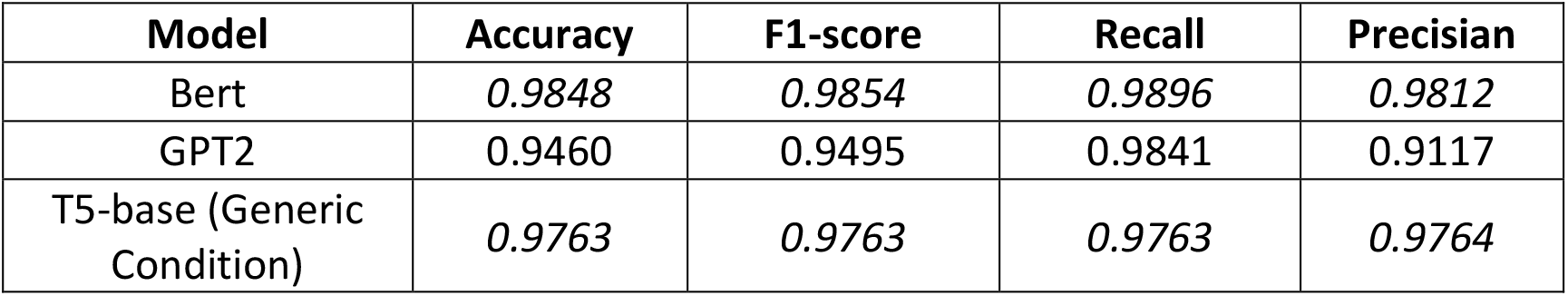
Performance metrics (Accuracy, F1-Score, Recall, and Precision) on the test dataset for three models: BERT-base, GPT-2, and T5-base.

| Model | Accuracy | F1-score | Recall | Precisian |
| --- | --- | --- | --- | --- |
| Bert | 0.9848 | 0.9854 | 0.9896 | 0.9812 |
| GPT2 | 0.9460 | 0.9495 | 0.9841 | 0.9117 |
| T5-base (Generic Condition) | 0.9763 | 0.9763 | 0.9763 | 0.9764 |

The confusion matrices in Figure 3 for BERT, GPT-2, and T5-base further illustrate the performance of these models, providing a detailed breakdown of true positives, true negatives, false positives, and false negatives, which helps in understanding their classification strengths and potential areas for improvement.

**Figure 3.**
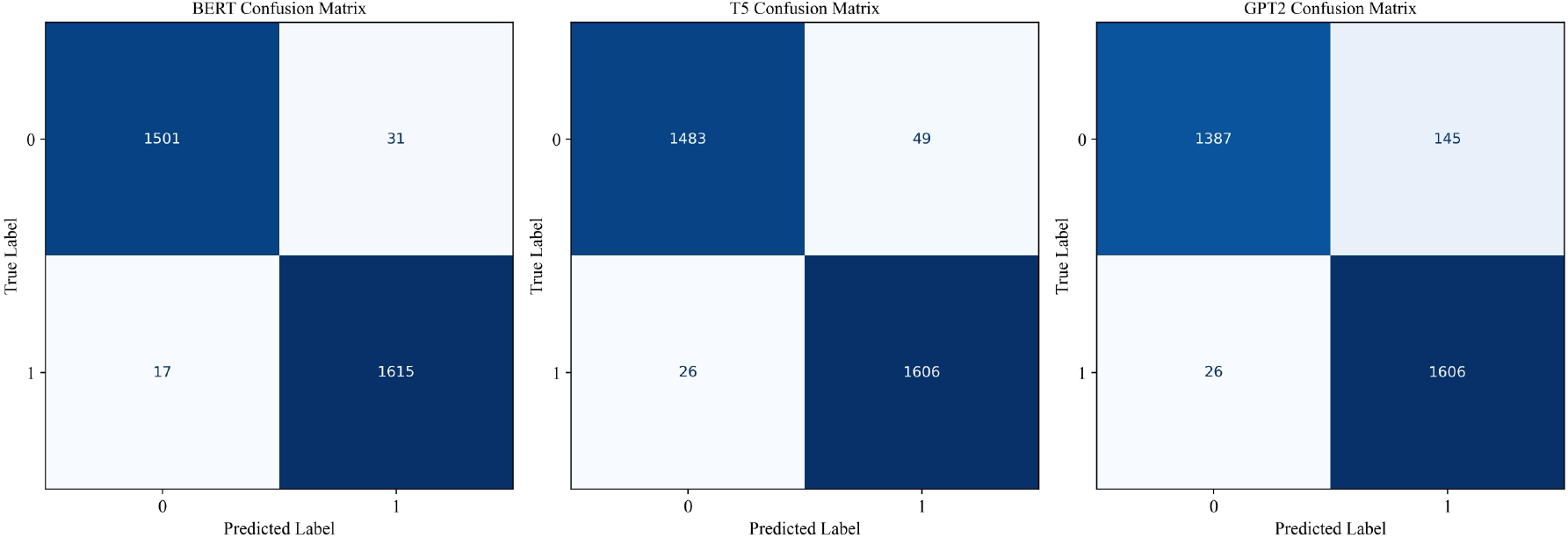
Confusion matrices illustrate the performance of the three binary classification models (BERT, GPT-2, and T5-base).

To evaluate the accuracy of our model on AI-generated data, we utilized the dataset provided in the study in (37). We tested our fine-tuned BERT model on this dataset, and the results are presented in Table 2. The findings indicate a significant reduction in the number of false positives (FP), decreasing from 27 to 15, representing a 44% improvement. Additionally, Figure 4 shows the confusion matrix for our fine-tuned BERT model when applied to the AI-generated misinformation dataset.

**Table 2.**
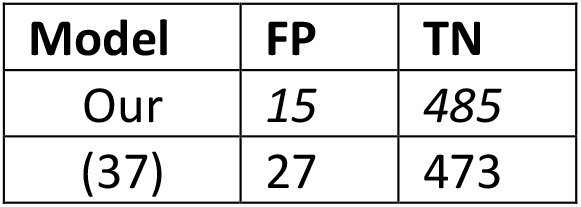
The table presents the False Positive (FP) and True Negative (TN) results obtained from testing the fine-tuned BERT model on our combined dataset, compared with the results reported in the (37) paper.

**Figure 4.**
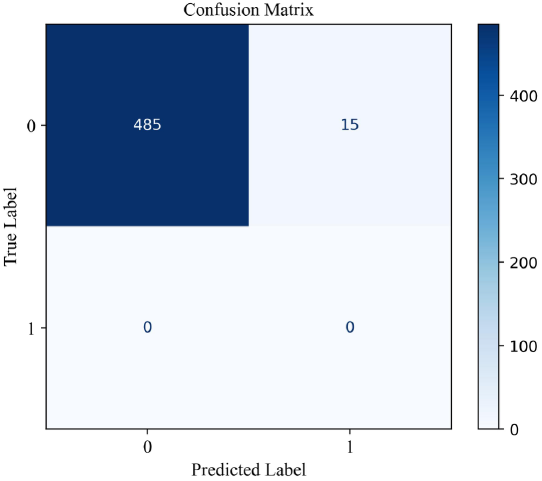
The figure displays the confusion matrix of our fine-tuned BERT model evaluated on the AI-generated dataset (37).

### Topic Models

We used the IFCN dataset, one of the largest datasets on COVID-19, to apply and evaluate topic modeling approaches. Three models were tested: LDA, Top2Vec, and BERTopic. After applying each topic model, the top 10 words associated with each topic were selected, and the three metrics—CV, NPMI, and IRBO—were computed to compare the models. (Table 3) summarizes the results across these metrics. Among the models, BERTopic achieved the highest scores across all metrics, leading to its selection for further analysis.

**Table 3.**
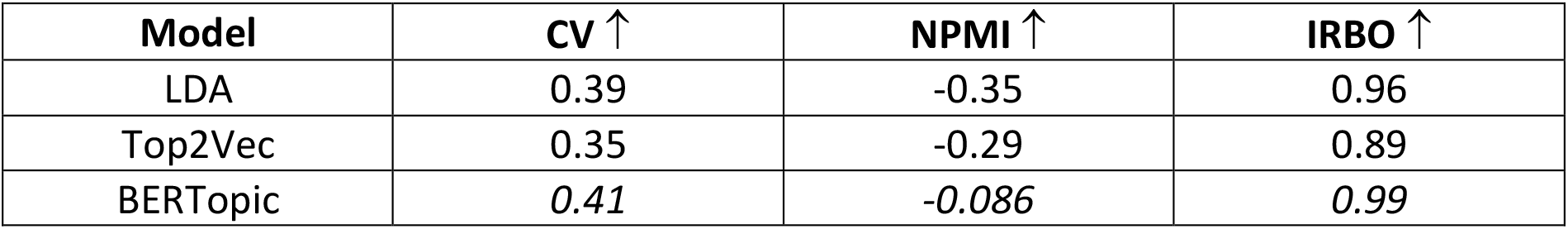
The table presents the performance of three topic modeling approaches—LDA, Top2Vec, and BERTopic— evaluated across three metrics: CV (Coherence Value), NPMI (Normalized Pointwise Mutual Information), and IRBO (Inverse Ranked Based Overlap). For all metrics, higher values indicate better performance.

| Model | CV ↑ | NPMI ↑ | IRBO ↑ |
| --- | --- | --- | --- |
| LDA | 0.39 | -0.35 | 0.96 |
| Top2Vec | 0.35 | -0.29 | 0.89 |
| BERTopic | 0.41 | -0.086 | 0.99 |

Many topic modeling approaches, including BERTopic, often encounter limitations when applied to real-world datasets, as they are unable to assign topics to all documents. This can leave a subset of documents unclassified, reducing the overall effectiveness of the model. To address this issue, we have developed Algorithm 1, a method for ensuring comprehensive topic assignment across the dataset.

This algorithm refines topic modeling by addressing unclassified documents after applying the BERTopic model. First, BERTopic is applied to a dataset of text documents, grouping those with similar semantic characteristics into distinct topics (Step 1). However, some documents may remain unclassified if they do not strongly align with any detected topic. To address this, all documents are converted into vector representations using a sentence transformer model, such as BERT embeddings, which capture their semantic meaning in a structured format (Step 2). To enhance processing efficiency, Uniform Manifold Approximation and Projection (UMAP) is then applied, reducing the dimensionality of document vectors while preserving essential semantic relationships (Step 3). Next, the algorithm computes the center of each topic cluster by averaging the vector representations of all documents assigned to that topic, with these centers serving as reference points that define the core meaning of each group (Step 4). For documents that remain unclassified, the algorithm determines the most appropriate topic assignment by computing cosine similarity between each unclassified document’s vector and the precomputed topic centers (Step 5). Finally, each unclassified document is assigned to the topic cluster whose center exhibits the highest similarity score, ensuring alignment with the most relevant category (Step 6).

This approach not only ensures that every document in the dataset is assigned a topic, but also enhances the interpretability and usability of the topic modeling results. By leveraging the semantic structure of the dataset, our algorithm effectively bridges the gap between unassigned documents and existing topic clusters, making it a robust solution for comprehensive topic coverage.

#### Algorithm 1 Assign a document to the closest topic

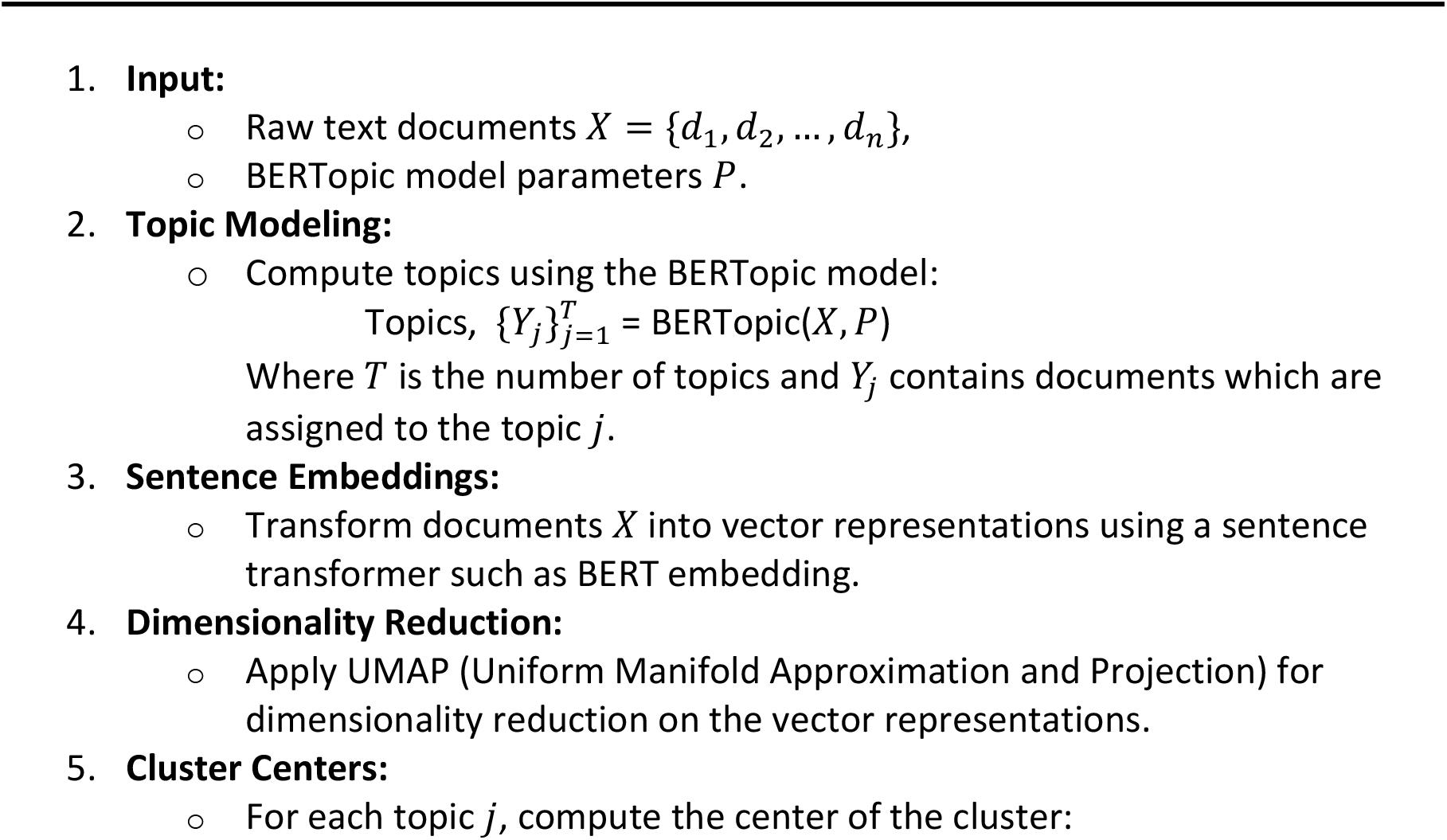
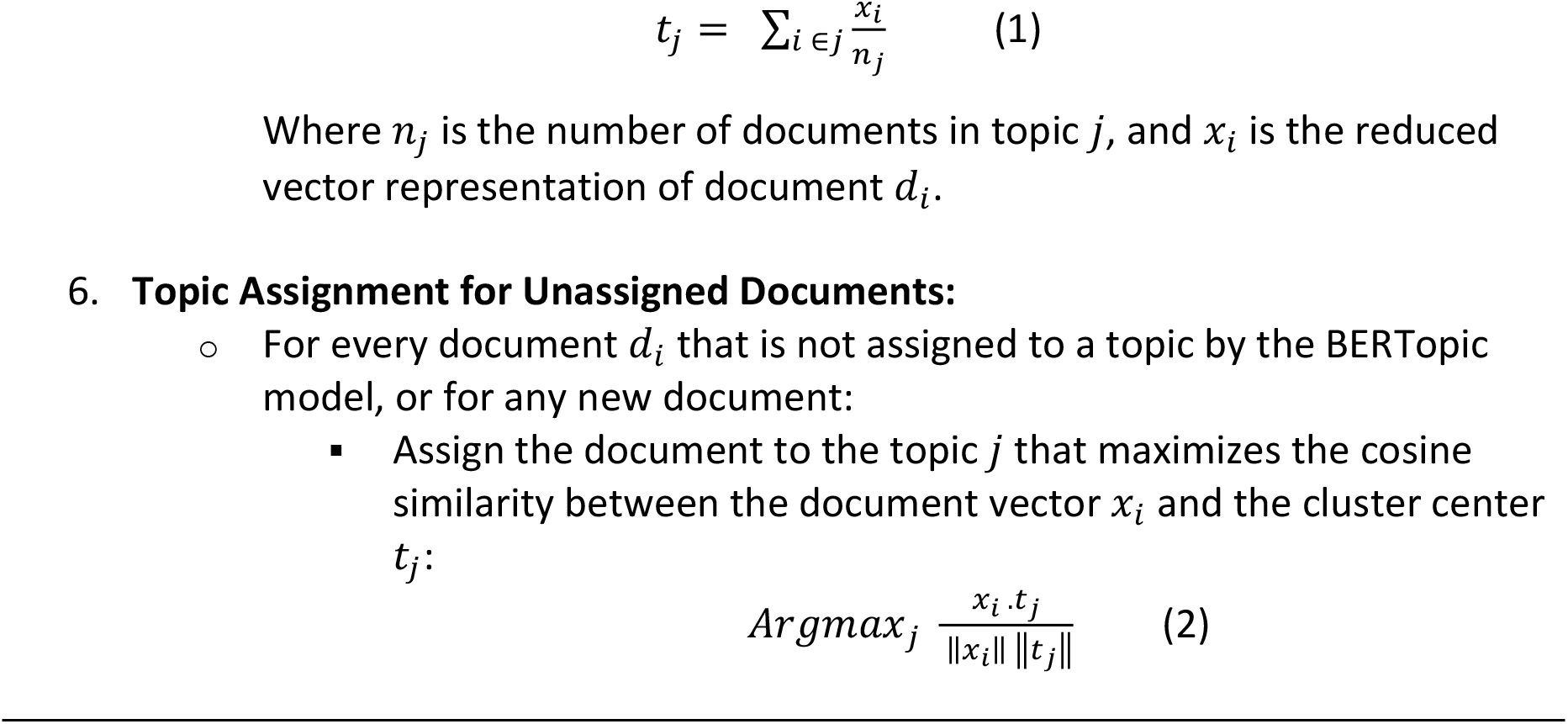

### Topic Description

As described in the Method section, topic modeling algorithms typically produce word-level representations of topics. While useful for identifying key terms associated with a topic, these representations often lack sufficient contextual information, making interpretation challenging. To overcome this limitation, we developed a structured prompt framework (outlined in Text box 1) and used the advanced capabilities of large language models (LLMs), specifically ChatGPT-4.0, to generate sentence-level representations for the identified topics. These sentence-level representations provide richer context and more interpretable descriptions, enabling a deeper understanding of the topics.

The prompt includes the top 10 most representative words for each topic as identified by the topic modeling algorithm and to add context and depth to the topic descriptions, we select five documents that are closest to the center of the corresponding topic cluster. The selection of these documents is guided by cosine similarity, performed using Equations 1 and 2, which measure the proximity of documents to the cluster center in the semantic space. An example of this process is provided in Text Box 1, illustrating how the top words and representative documents are integrated into the prompt to produce a high-quality sentence-level representation.

By combining these elements, we construct detailed and context-rich prompts that guide ChatGPT-4.0 in generating coherent and semantically accurate sentence-level topic representations. This approach ensures that the abstract themes identified by topic modeling are translated into human-readable and interpretable descriptions.

#### Text box 1

*The prompt structure and one example to find topics description mentioned in the text box*.

**Topic Description Prompt Structure:**

System Role:

> **Topic main words:** [Top 10 words]
>
> **Topic document examples:** [5 closest examples to the center of the topic]

User Role:

> **“Describe topic in a short phrase?”**

**Topic Description Prompt Example:**

System Role:

> **Topic main words:** [‘masks’, ‘mask’, ‘face’, ‘wearing’, ‘wear’, ‘use’, ‘oxygen’, ‘hypoxia’, ‘cause’, ‘you’].
>
> **Topic document examples:**
>
> 1. CDC does not recommend wearing masks.
> 2. The US Centers for Disease Control and Prevention (CDC) contradicted itself by advising people to wear cloth masks against the novel coronavirus while also saying masks do not stop smoke inhalation during a wildfire.
> 3. The WHO changed its mind about masks and now says that they can increase the risk of infection.
> 4. Non-medical masks are ineffective in preventing the spread of the disease are circulating online.
> 5. Whether CDC had scheduled announcement that all should wear masks for everyday life.

User Role: “**Describe topic in a short phrase?”**

Output Answer: **“Controversies and Debates over Mask Wearing and its Effectiveness”**

To evaluate the quality of the generated topic descriptions, we engaged three independent raters to assess the descriptions based on three categories: appropriate, somewhat appropriate, and not appropriate. The evaluation results are presented in Table 4 and highlights that the majority of topic descriptions were well-received. Specifically, the total proportion of accepted descriptions (the sum of those rated as appropriate and somewhat appropriate) was 99.6%. This acceptance rate demonstrates the effectiveness and reliability of the proposed method for generating meaningful and contextually relevant topic descriptions.

**Table 4.**
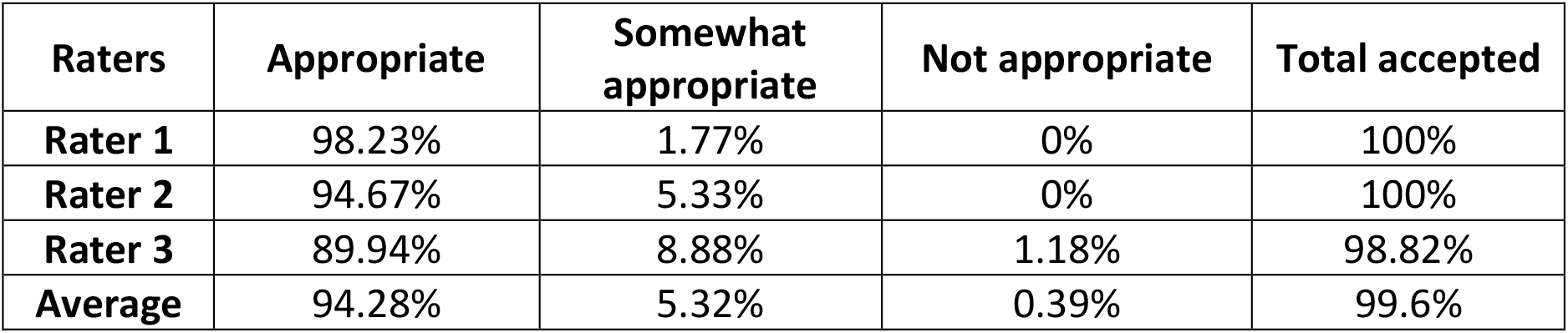
The table presents the percentage of topic descriptions rated as “Appropriate,” “Somewhat Appropriate,” and “Not Appropriate” by each rater, along with the total number of accepted topic descriptions, calculated as the sum of those rated “Appropriate” and “Somewhat Appropriate”.

| Raters | Appropriate | Somewhat appropriate | Not appropriate | Total accepted |
| --- | --- | --- | --- | --- |
| **Rater 1** | 98.23% | 1.77% | 0% | 100% |
| **Rater 2** | 94.67% | 5.33% | 0% | 100% |
| **Rater 3** | 89.94% | 8.88% | 1.18% | 98.82% |
| **Average** | 94.28% | 5.32% | 0.39% | 99.6% |

After generating concise descriptions for each topic, we used the structured prompt framework outlined in Text box 2, which includes a list of these topic descriptions. This structured prompt was then input into the ChatGPT-4.0 API to further refine and categorize the topics into overarching themes.

The output from this process not only provides a clear categorization of topics into distinct themes but also includes a concise description for each theme. This step ensures that the topics are grouped in a meaningful and interpretable way, facilitating a deeper understanding of the data’s thematic structure.

The categorized topics and their corresponding theme descriptions are provided in Figure 5 and Figure 6, showcasing the effectiveness of the proposed method in generating coherent and insightful thematic groupings.

**Figure 5.**
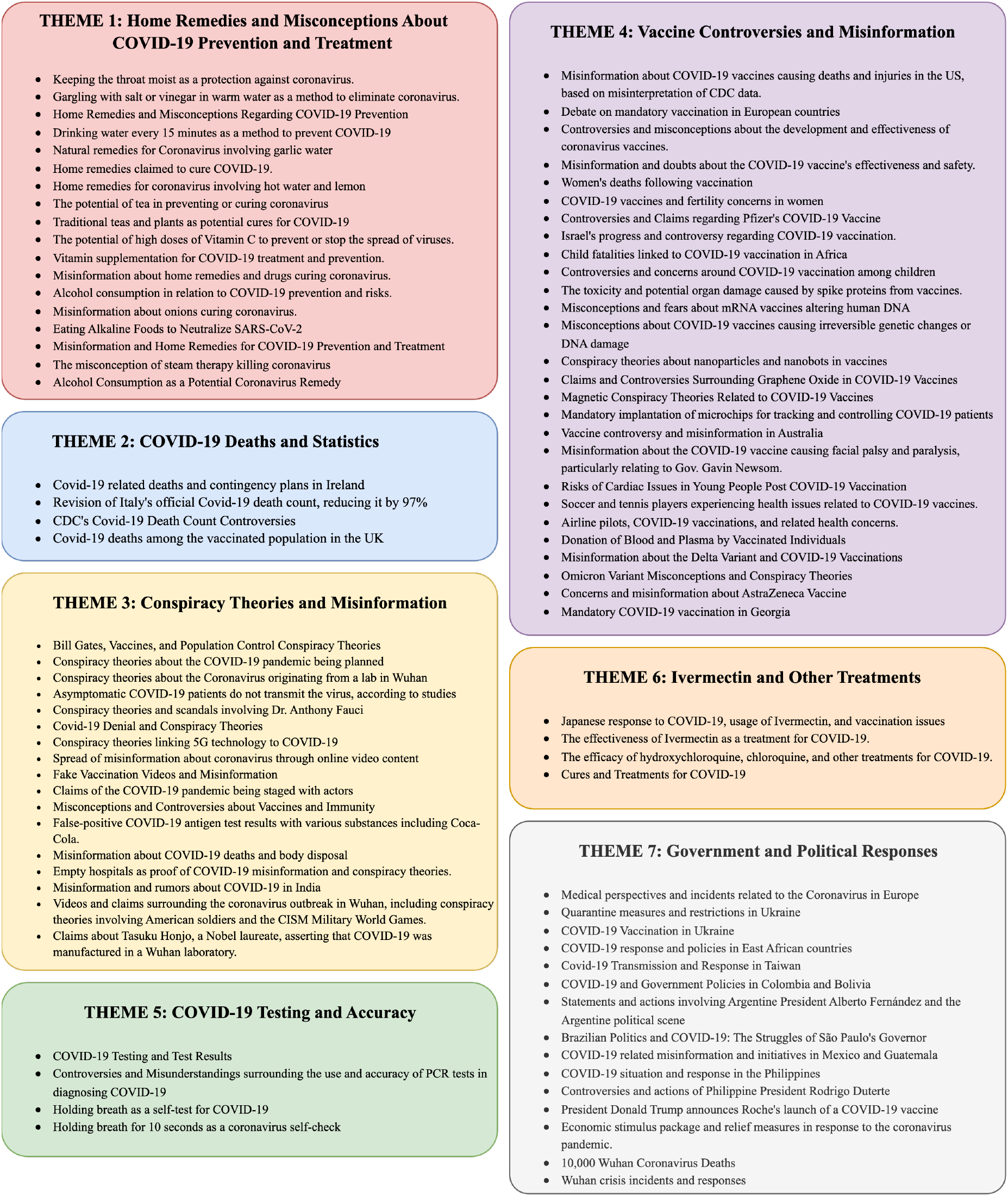
Descriptions of Themes 1 to 7 along with the corresponding topic descriptions assigned to each theme.

**Figure 6.**
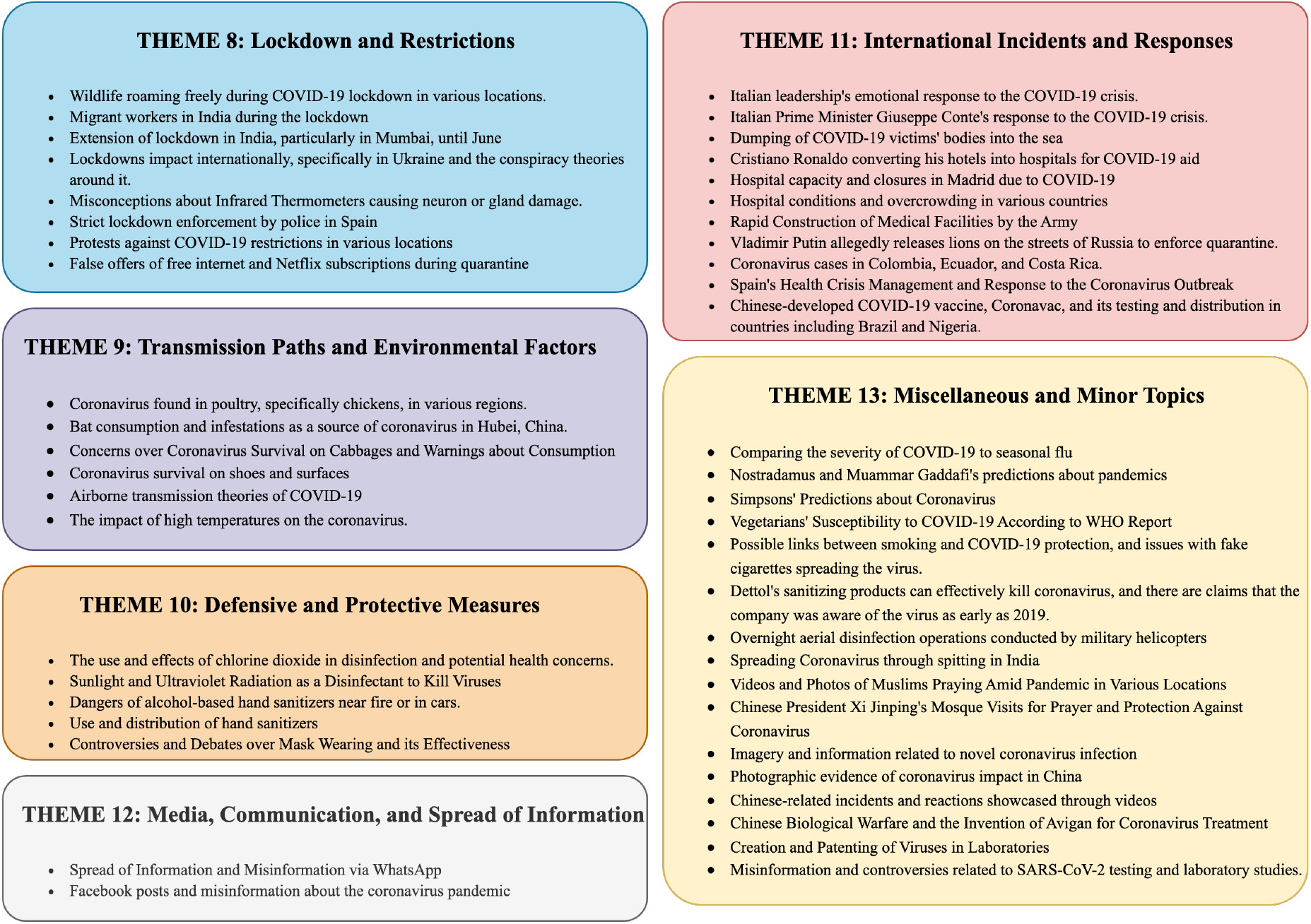
Descriptions of Themes 8 to 13 along with the corresponding topic descriptions assigned to each theme.

#### Text box 2

*Structure of the prompt for identifying topic themes*.

**Finding Topic Themes Prompt Structure:**

System Role:

> **The following are topics related to COVID-19. Go through all topics and categorize them into relevant groups. Mention topics number for each category**.

User Role:

> **Topics description list**

Using Algorithm 1, we assign each document to a topic, enabling us to determine the distribution of each theme. Figure 7 shows the distribution of all themes, while Figure 8 shows the distribution of each topic within Theme 8 over time.

**Figure 7.**
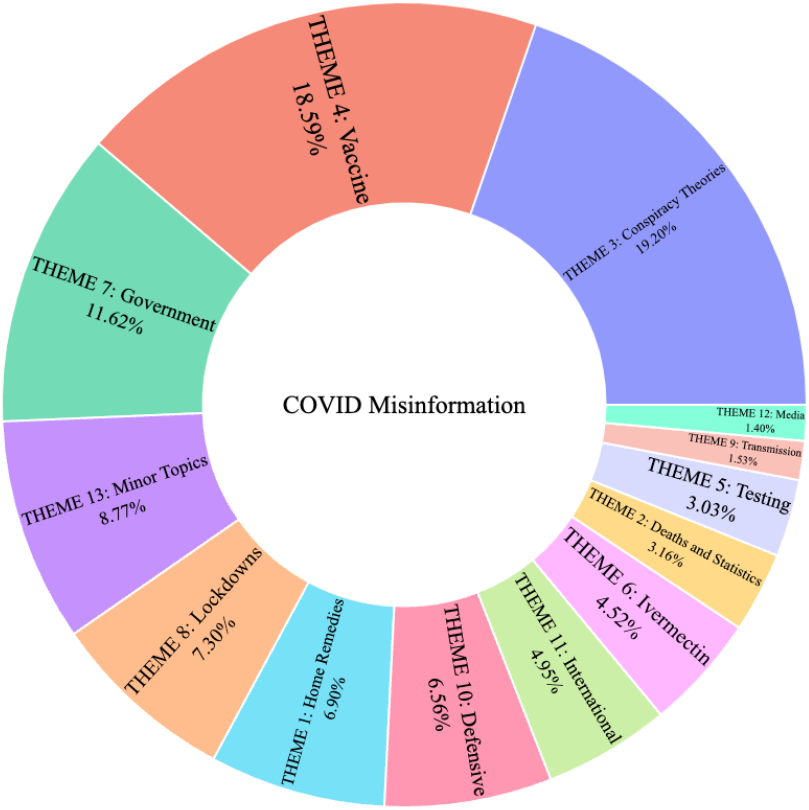
Distribution of COVID-19 misinformation themes.

**Figure 8.**
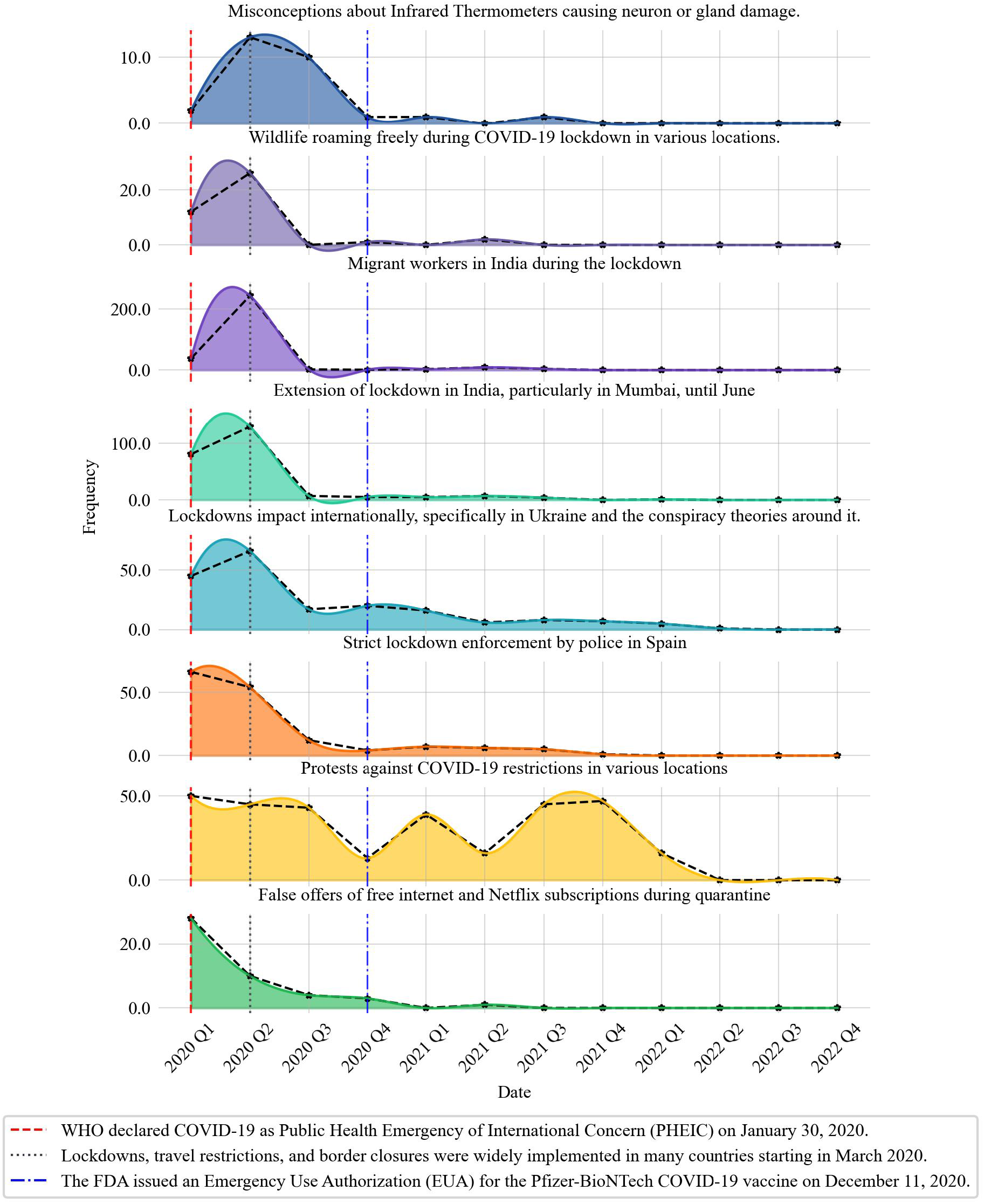
Distribution of THEME 8 topics during the time.

After identifying the misinformation themes, we leverage the explanations provided in the IFCN dataset as a basis to draw refutation arguments to address these themes. For each identified theme, we develop an refutation that aligns with its context, aiming to clarify the nature of the misinformation, its potential origins, and its impact. These refutations are presented in Text box 3- The refutation arguments list for 13 themes., providing valuable insights into the underlying reasons for the misinformation.

#### Text box 3

*The refutation arguments list for 13 themes*.

**Refutation List:**

1. Home remedies for COVID-19 prevention and treatment are often based on misinformation. While they may promote general health, no scientific evidence supports their effectiveness in preventing or curing COVID-19.
2. Misinformation about COVID-19 deaths, statistics, and their relation to vaccination often involves data misrepresentation, misinterpretation of health reports, or fabricated claims. Such falsehoods can cause confusion, erode trust in public health institutions, and foster skepticism about the impact of COVID-19 and the effectiveness of vaccination.
3. Conspiracy theories and misinformation related to COVID-19 often rely on unverified claims, distorted facts, or manipulated content. These falsehoods exploit fear, uncertainty, and distrust to spread misleading narratives about the pandemic’s origins, vaccines, and public health measures. Common conspiracy themes include claims of the pandemic being planned, unfounded accusations against prominent figures like Bill Gates, misinformation linking 5G technology to COVID-19, and fabricated stories about vaccines and immunity.
4. Misinformation about COVID-19 vaccines, including false claims about safety, efficacy, and side effects, has fueled skepticism and vaccine hesitancy worldwide. These falsehoods undermine public trust in vaccines.
5. Misinformation about COVID-19 testing often targets the accuracy of PCR tests and promotes unscientific self-check methods like holding one’s breath. PCR tests are highly accurate but may detect non-infectious viral fragments, which can be misunderstood. In reality, PCR tests are highly sensitive and accurate for detecting COVID-19 when used appropriately, though they may detect non-infectious viral fragments in some cases.
6. Misinformation about treatments for COVID-19, including the use of Ivermectin, hydroxychloroquine, and unproven supplements like Quermax, often exaggerate the effectiveness of these treatments or misrepresent scientific studies to support their use over proven measures like vaccines. While some drugs, such as Ivermectin and hydroxychloroquine, were initially studied for potential COVID-19 treatment, clinical trials have not provided sufficient evidence to confirm their safety or efficacy for this purpose. Similarly, supplements and alternative remedies lack the rigorous scientific validation required to prove their effectiveness against COVID-19.
7. Misinformation about government and political responses to COVID-19 involves distorted facts, fabricated claims, or misrepresentation of policies and actions. These falsehoods range from exaggerated accomplishments to unfounded accusations, including misreporting financial expenditures, misattributing statements to political leaders, or fabricating praise from international organizations. Governments worldwide have implemented a wide range of measures to combat COVID-19, including quarantine protocols, vaccination drives, economic relief packages, and public health campaigns. However, the interpretation and communication of these measures can vary, leading to the spread of misleading narratives. To counter this, it is crucial to verify information from credible sources, such as official government announcements, reputable media outlets.
8. Misinformation about lockdowns and COVID-19 restrictions often includes fabricated or misrepresented events, videos, or claims. False narratives range from altered timelines of events to conspiracies about government actions, protests, and public compliance. Lockdowns and restrictions were implemented globally to curb the spread of COVID-19, with varying levels of enforcement and public response. To combat misinformation, it is critical to verify claims through official government announcements, trusted media sources, and independent fact-checking organizations.
9. Misunderstandings about how COVID-19 spreads and survives in various environments have led to numerous false claims and fears. These include unverified assertions about the virus being linked to specific foods like chickens or cabbages, its survival on surfaces like shoes or roads, and exaggerated theories about airborne transmission and environmental factors like temperature. Scientific evidence indicates that while COVID-19 can survive on surfaces for varying durations, its primary transmission occurs through respiratory droplets and, in some cases, aerosols. Claims about the virus thriving on specific foods or being destroyed at particular temperatures lack scientific support.
10. Misleading or false claims about defensive and protective measures against COVID-19 have contributed to confusion and unsafe practices. These include exaggerated claims about the use of chlorine dioxide, UV rays, and alcohol-based products, as well as debates over the effectiveness of masks and sanitizers. While certain measures, such as wearing masks and using hand sanitizers, are scientifically proven to reduce the spread of the virus, other claims, like sunlight killing COVID-19 or chlorine dioxide being safe for disinfection, lack credible evidence or are outright false. Relying on guidance from reputable health organizations like the World Health Organization (WHO) and the Centers for Disease Control and Prevention (CDC) is essential for understanding which protective measures are effective and safe.
11. International Misinformation about international incidents and responses to COVID-19 involves exaggerated or fabricated claims about government actions, healthcare capacity, or emotional reactions of leaders. Examples include false reports of rapid hospital construction, misattributed emotional responses by world leaders, or unverified claims about the treatment of COVID-19 victims. Such false narratives misrepresent the global efforts to combat the pandemic and can erode trust in public health measures and international cooperation. Misleading claims, such as countries discarding victims’ bodies at sea or building thousands of hospital beds in days, often stem from misinterpreted images or old videos unrelated to COVID-19. To combat misinformation, it is essential to rely on verified data from credible sources like independent fact-checking organizations.
12. Misinformation about COVID-19 spreads rapidly through social media platforms and messaging apps like WhatsApp and Facebook. False claims often exploit fear and uncertainty, such as fabricated government policies, misleading features of apps, or unfounded accusations about media manipulation. To counter this, it is crucial to verify claims through reliable fact-checking organizations and official communications.
13. These claims lack credible scientific evidence or are distortions of real information, such as the exaggerated impact of products, religious practices, or fabricated connections to bioweapons and patented viruses. While some of these topics may seem minor or humorous, they can still misinform the public and detract from accurate understandings of the pandemic. Critical thinking, fact-checking, and reliance on credible scientific and official sources are vital to combatting these misinformation.

### Provide Refutation

In the final stage of our process, we design a prompt text to enable ChatGPT-4.0 to detect specific misinformation themes. The prompt text includes a detailed description of the themes and a question-answer list. To create this question-answer list, we select the document closest to the center of each topic and associate it with the corresponding theme. Detailed information about the prompt text can be found in Text box 4.

#### Text box 4

*Prompt structure and one example to find a document theme*.

**Finding Document Themes Prompt Structure:**

System Role:

> **“The following is the description of topic themes related to COVID-19 misinformation. Find the closest theme for the given text. Answer in a consistent style.”**

User Role:

> **Themes description list**

Assistant Role:

> **Question-Answer List**

User Role:

> **Input text**

**Finding Topic Themes Prompt Example:**

System Role:

> **“The following is the description of topic themes related to COVID-19 misinformation. Find the closest theme for the given text. Answer in a consistent style.”**

User Role:

> **Themes description list**

Assistant Role:

> **Question-Answer List**

User Role:

> **“A video shows that Bill Gates admits the vaccine will no doubt kill 700**,**000 people”**

Output Answer:

> **THEME 3: “Conspiracy Theories and Misinformation”**

To evaluate our approach, we randomly select 50 documents from the IFCN dataset that are not included in the question-answer list. We then tested the prompting method with ChatGPT-4.0, achieving an 80% accuracy rate in detecting the correct themes as shown in Table 5.

**Table 5.**
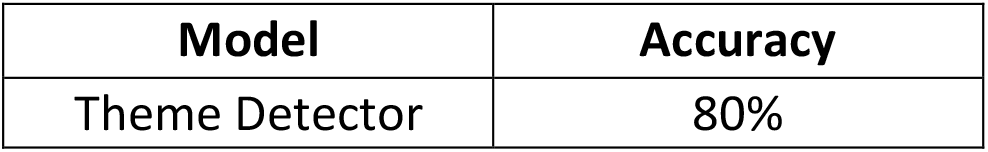
Accuracy of the model for detection document’s theme.

| Model | Accuracy |
| --- | --- |
| Theme Detector | 80% |

### Proposed System

Based on the process we introduced, we propose the development of a Misinformation Detection and Inoculation System (MDIS). This system is designed to first determine whether a given text document contains misinformation or not. If the document is identified as containing misinformation, the system then detects its theme and provides a detailed refutation of the misinformation. The primary objectives of MDIS are to prevent the spread of misinformation and to enhance public health knowledge.

MDIS operates by integrating three key components. First, it utilizes a trained large language model (LLM) to classify text documents as either misinformation or non-misinformation. Next, it employs another trained LLM to detect the specific misinformation theme within documents identified as containing misinformation. Finally, it leverages a refutation list, which is generated during the theme description phase, to provide context and counter-narratives for each detected theme. This comprehensive approach enables the system to effectively address misinformation while equipping users with accurate information.

Figure 9 illustrates an example of an input to MDIS and its corresponding output, demonstrating how the system analyzes a document, identifies misinformation, detects the associated theme, and presents an explanatory response.

**Figure 9.**
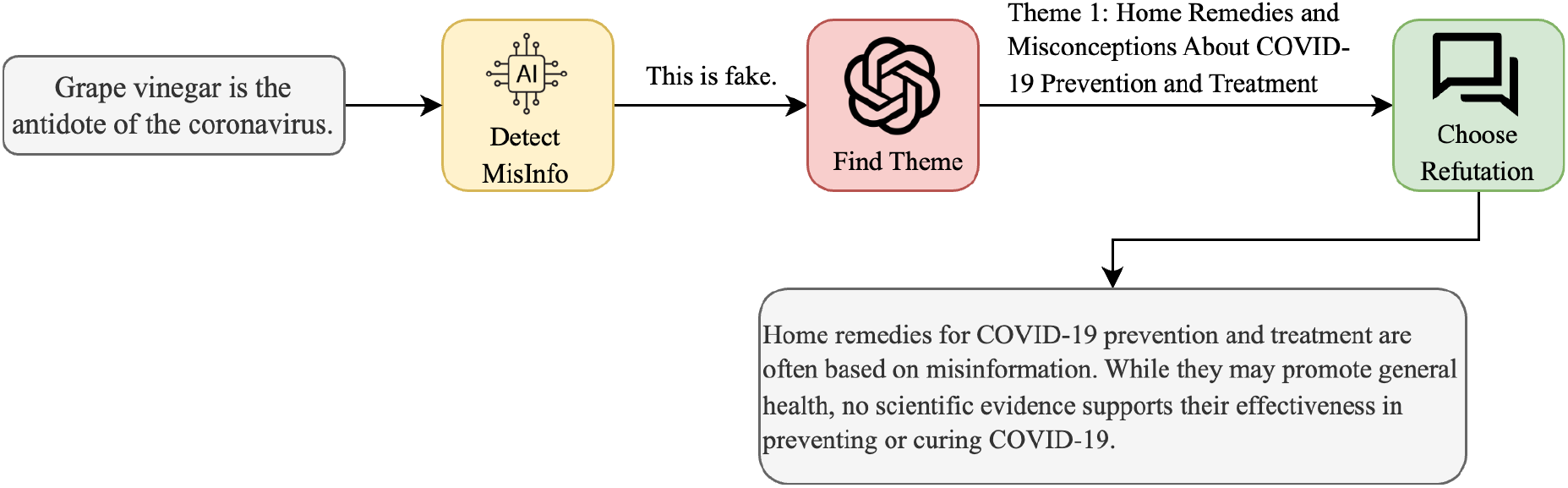
One input text example and the output of Misinformation Detection and Inoculation System (MDIS).

## Discussion

### Principle Results

In this study, we developed a Misinformation Detection and Inoculation Process (MDIP), which is designed to analyze the dynamics of misinformation and facilitate the creation of a Misinformation Detection and Inoculation System (MDIS). MDIS provides detailed refutations for misinformation circulating on social media, helping to combat its spread and improve public understanding. MDIP consists of four main steps (subroutines), each contributing to the system’s overall effectiveness.

The first step focuses on Detecting Misinformation. In this phase, we trained three large language models (LLMs) to classify text documents as misinformation or non-misinformation. Among these, the BERT model achieved the highest accuracy of 98%. Additionally, we demonstrated that enriching the training dataset with both formal and informal language significantly improved the system’s ability to identify AI-generated misinformation, reducing false positives (FP) by 44% compared to previous models. This enrichment ensures the model performs well across various linguistic styles and sources.

The second step involves Identifying Misinformation Topics. For this, we used the IFCN dataset, one of the most comprehensive collections of COVID-19-related misinformation. We applied three topic modeling techniques—LDA, Top2Vec, and BERTopic—to analyze the dataset. Among these, BERTopic outperformed the others, achieving the highest scores in CV at 0.41, NPMI at -0.086, and IRBO at 0.99. Once the optimal topic model was identified, we developed Algorithm 1 to assign documents to their closest topics. Since most topic models often leave outliers unassigned, Algorithm 1 ensures that every document is categorized under a topic, enabling more comprehensive topic distribution analysis.

In the third step, we focused on deriving Sentence-Level Representations for Topics. While most topic modeling approaches provide word-level representations, we extended this by developing sentence-level representations for each topic. These representations were then used to identify broader themes within the misinformation topics. Based on these themes, we created detailed refutations to address the key elements and nuances of the misinformation being analyzed.

The fourth step involved designing a prompt text for theme detection using ChatGPT-4.0. This prompt was crafted to detect the theme of a misinformation document accurately. By integrating the themes and refutations, our prompt-based theme detector achieved an accuracy of 80%, demonstrating its effectiveness in identifying misinformation themes.

Finally, by combining the misinformation detector, theme detector, and refutation generator, we developed the Misinformation Detection and Inoculation System (MDIS). This system can accurately identify misinformation, determine its underlying theme, and provide a suitable refutation to counteract its spread. MDIS represents a comprehensive tool for combating misinformation and enhancing public awareness in the digital age.

### Limitations

While this study presents a robust framework for detecting and addressing misinformation, several limitations should be acknowledged. First, the system’s performance relies on the quality and diversity of the training datasets, which may not fully capture the linguistic and contextual nuances of misinformation in different regions, languages, or cultural contexts. The model has been tested solely on COVID-19-related misinformation, and its effectiveness for other domains remains unverified. Additionally, it has not been evaluated for multilingual adaptation, meaning its ability to detect misinformation across different languages and sociocultural contexts remains uncertain. Moreover, while the theme detection module achieved 80% accuracy, its accuracy could be further improved, particularly in handling ambiguous or overlapping themes. False positives and negatives remain a concern, particularly when misinformation contains opinion-based, satirical, or context-dependent elements. Additionally, the refutations generated by the system, though informative, might require additional refinement to ensure they are universally comprehensible and contextually appropriate for diverse audiences. While these refutations aim to enhance transparency, their AI-generated nature may not always align with human reasoning or ethical considerations.

Additionally, the system has not yet been extensively tested in real-world applications, which limits understanding of its practical impact on misinformation spread and public health outcomes. Furthermore, misinformation evolves over time, and a model trained on past narratives may require periodic retraining to remain effective against emerging falsehoods, including AI-generated misinformation. Finally, the reliance on automated methods raises potential concerns about interpretability and transparency, which are crucial for fostering trust and adoption by end users.

### Comparison with Prior Work

The proposed Misinformation Detection and Inoculation Process (MDIP) and the resulting Misinformation Detection and Inoculation System (MDIS) build upon and advance the body of research focused on misinformation detection and mitigation. Previous research has demonstrated the efficacy of machine learning (ML) models, particularly deep learning approaches, in detecting misinformation. They employed ML techniques to classify fake news using textual features, demonstrating the value of automated detection methods (50,53,68). Our study extends these efforts by integrating enriched datasets containing both formal and informal language styles, ensuring better generalization across diverse linguistic sources, including AI-generated misinformation.

Topic modeling techniques such as LDA have been used in prior studies to analyze misinformation (39,57,69). Our approach improves on these works by addressing limitations in document assignment and theme interpretation. We developed an algorithm to assign every document to the most relevant topic, resolving the common issue of unclassified documents in topic modeling. Additionally, we moved beyond word-level topic representations to generate sentence-level descriptions, offering richer and more interpretable insights. Finally, we designed an effective prompt text to automatically identify the themes of misinformation. This automated approach reduces reliance on manual interpretation, minimizing human bias and increasing scalability.

Many prior studies have addressed misinformation detection or topic analysis in isolation. They analyzed misinformation using sentiment analysis but did not integrate detection with thematic analysis and did not provide a framework for counteracting misinformation (39,70,71). Our work unifies detection, topic modeling, thematic refutation, and public health intervention in a single framework. The MDIS framework automates the end-to-end process, offering a scalable solution to tackle the complexity of misinformation dynamics.

## Conclusions

This study introduces a novel framework for addressing health-related misinformation through the development of the Misinformation Detection and Inoculation Process (MDIP) and the Misinformation Detection and Inoculation System (MDIS). By leveraging advanced natural language processing techniques, MDIS provides an automated approach to detecting misinformation, identifying its underlying themes, and delivering clear and contextually relevant refutations. This approach is designed to combat the rapid spread of misinformation on social media and improve public health knowledge.

The integration of enriched datasets ensures the system can handle diverse linguistic styles, making it adaptable to various contexts and sources of misinformation. Additionally, the ability to generate sentence-level representations and thematic refutations allows for a deeper understanding of misinformation dynamics and provides actionable insights to counter false narratives effectively.

MDIS represents a scalable and comprehensive solution to the misinformation problem, addressing both its detection and mitigation in a unified framework. While challenges remain, such as adapting to regional and cultural contexts and improving thematic interpretation, the system sets the stage for future advancements in misinformation management.

In conclusion, this work provides a robust and intelligent tool for combating misinformation, contributing to the broader goal of promoting health literacy and trust in credible information sources. By equipping individuals and public health authorities with an effective system to manage misinformation, this framework offers significant potential to improve public health communication and outcomes in an increasingly digital world.

## Data Availability

The study used only openly available human data that were publicly accessible prior to the initiation of the study. These datasets were originally located at:
1- AAAI 2021 COVID-19 Fake News Dataset:
https://github.com/diptamath/covid_fake_news
2- COVID-19FNIR Dataset:
https://ieee-dataport.org/open-access/covid-19-fake-news-infodemic-research-dataset-covid19-fnir-dataset
3- IFCN Fact-Checked Claims:
https://www.poynter.org/ifcn-covid-19-misinformation/

https://ieee-dataport.org/open-access/covid-19-fake-news-infodemic-research-dataset-covid19-fnir-dataset

https://www.poynter.org/ifcn-covid-19-misinformation/

## Acknowledgements

This research is supported in part by a research grant from the Investigator Initiated Studies Program of Merck Sharp & Dohme Corp (MISP #102050). The opinions expressed in this paper are those of the authors and do not necessarily represent those of Merck Sharp & Dohme Corp.

## Ethics Statement

This study did not involve human subjects, human tissue, or identifiable private information.. All data analyzed in this study were obtained from publicly available, de-identified sources, including the AAAI 2021 Competition Dataset, the COVID-19FNIR dataset, and the IFCN COVID-19 fact-checking corpusThe research complied with all relevant ethical standards and data use policies, and poses no risk to individuals or communities. The study’s sole objective is to advance computational methods for understanding and mitigating the spread of health misinformation.

## Abbreviations

JMIR: Journal of Medical Internet Research
LLM: Large Language Models
WHO: World Health Organization
CDC: Centers for Disease Control and Prevention
ML: Machine Learning
LDA: Latent Dirichlet Allocation
CV: Coherence Value
NPMI: Normalized Pointwise Mutual Information
RBO: Rank Biased Overlap
IRBO: Inverted RBO
HPV: Human Papillomavirus
MDIP: Misinformation Detection and Inoculation Process
MDIS: Misinformation Detection and Inoculation System
DNN: Deep Neural Network
AI: Artificial Intelligence
BERT: Bidirectional Encoder Representations from Transformers

